# Immune boosting bridges leaky and polarized vaccination models

**DOI:** 10.1101/2023.07.14.23292670

**Authors:** Sang Woo Park, Michael Li, C. Jessica E. Metcalf, Bryan T. Grenfell, Jonathan Dushoff

## Abstract

Two different epidemiological models of vaccination are commonly used in dynamical modeling studies. The leaky vaccination model assumes that all vaccinated individuals experience a reduced force of infection by the same amount. The polarized vaccination model assumes that some fraction of vaccinated individuals are completely protected, while the remaining fraction remains completely susceptible; this seemingly extreme assumption causes the polarized model to always predict lower final epidemic size than the leaky model under the same vaccine efficacy. However, the leaky model also makes an implicit, unrealistic assumption: vaccinated individuals who are exposed to infection but not infected remain just as susceptible as they were prior to exposures (i.e., independent of previous exposures). To resolve the independence assumption, we introduce an immune boosting mechanism, through which vaccinated, yet susceptible, individuals can gain protection without developing a transmissible infection. The boosting model further predicts identical epidemic dynamics as the polarized vaccination model, thereby bridging the differences between two models. We further develop a generalized vaccination model to explore how the assumptions of immunity affect epidemic dynamics and estimates of vaccine effectiveness.

**Significance statement:** Different assumptions about the long- and medium-term effects of protective vaccination can predict sharply different epidemiological dynamics. However, there has been limited discussion about which assumptions are more realistic and therefore more appropriate for making public health decisions. Here, we show that the differences between the two most common assumptions (the “leaky” and “polarized” vaccination models) are bridged by immune boosting, a mechanism by which individuals who resist infectious challenge due to partial immunity have their immunity increased. We demonstrate that this mechanism has important implications for measuring vaccine effectiveness. Our study challenges fundamental assumptions about commonly used vaccination models and provides a novel framework for understanding the epidemiological impact of vaccination.

## Introduction

Vaccination plays a critical role in controlling infectious disease outbreaks by protecting against new infections and associated disease (Iwasaki and Omer, 2020). In particular, if a critical vaccination threshold is reached, the reproduction number (defined as the average number of secondary infections caused by an infected individual) is reduced to below 1, and future epidemics can be prevented (Anderson and May, 1985). But reaching a critical vaccination threshold can be challenging, and vaccines often provide imperfect protections (Gandon et al., 2003; Anderson et al., 2020).

There are two main ways of modeling vaccines with imperfect protections: “leaky” and “all-or-nothing” vaccine (Smith et al., 1984). The leaky vaccination model assumes that vaccinated individuals experience a reduced force of infection (e.g., multiplied by a factor 1 − VE_L_ *<* 1). The “all-or-nothing” vaccination model assumes that the proportion VE_P_ of vaccinated individuals are completely protected and the remaining proportion 1 − VE_P_ of vaccinated individuals are completely susceptible. This model is analogous to the polarized immunity model, in which infection from one strain gives complete or no protection against other strains (Gog and Grenfell, 2002)—we thus refer to this model as the polarized vaccination model (Gomes et al., 2014). Here, both VE_L_ and VE_P_ represent vaccine efficacy, which we define as the proportion of people protected from their first challenge.

When these two models are used with the same nominal vaccine efficacy VE_L_ = VE_P_, they predict different epidemic dynamics, including the final size (Smith et al., 1984): for high force of infection, almost all individuals eventually get infected in the leaky model, whereas many individuals are permanently protected in the polarized model. Modelers tend to rely on the leaky assumption, including throughout the SARS-CoV-2 pandemic (Dyson et al., 2021; Gozzi et al., 2021; Marziano et al., 2021; Matrajt et al., 2021; Park et al., 2022) with some exceptions (Bubar et al., 2021; Buckner et al., 2021). Various reasons have been given, but most likely is a combination of convenience and tradition.

Both models represent simplifications of reality. The leaky model in particular overlooks a potentially important mechanism: individuals in this model do not lose any susceptibility when (implicitly) exposed to a challenge that does not result in infection. In fact, vaccinated individuals who successfully fight off exposures can experience immune boosting, thus becoming less susceptible to future infections without becoming infectious or developing symptoms from the exposure (Lavine et al., 2011; Yang et al., 2020).

In this study, we compare different approaches to dynamical modeling of vaccination and immunity. First, we construct a model with leaky vaccination and boosting, and show that the transmission dynamics of this model can bridge from the dynamics of the standard leaky model (with no boosting) to those of the polarized model (with perfect boosting). Then, we construct a generalized vaccination model, which includes all three mechanisms, and explore its dynamics. Finally, we use our framework to compare measures of vaccine efficacy.

## Mathematical models of vaccine-induced immunity

Throughout the paper, we assume that a population mixes homogeneously and that there is no loss of immunity; the latter assumption essentially corresponds to focusing on a single outbreak. We begin with a standard SIR model with a leaky vaccine, in which all vaccinated individuals experience a reduced force of infection by a factor of 1 − VE_L_:

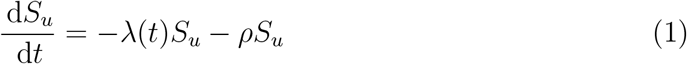

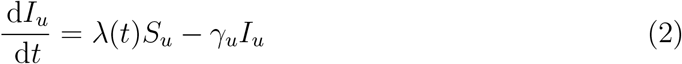

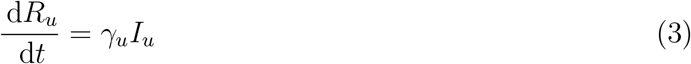

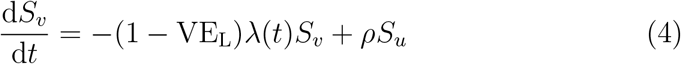

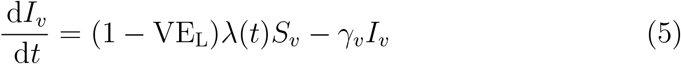

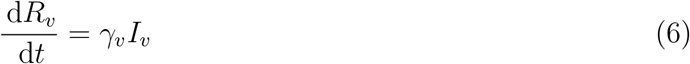

where subscripts *u* and *v* indicate the unvaccinated and vaccinated individuals; *λ* represents the baseline force of infection experienced by unvaccinated individuals; *ρ* represents vaccination rate; *γ* represents the recovery rate; and VE_L_ represents the vaccine efficacy, which also captures the amount of reduction in the probability of infection. This kind of model is sometimes called “history-based”, since susceptibility of an individual depends only on their history of infections (or vaccination) (Gog and Grenfell, 2002; Gog and Swinton, 2002; Kucharski et al., 2016).

Conversely, the polarized vaccination model assumes that a proportion VE_P_ of vaccinated individuals become fully immune, whereas the remaining proportion 1 − VE_P_ remain susceptible:

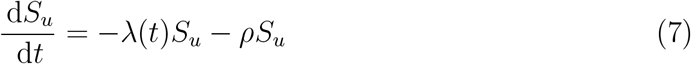

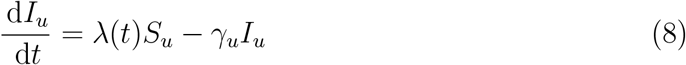

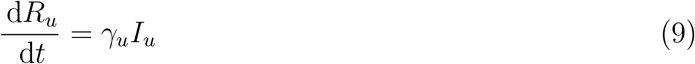

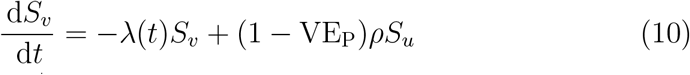

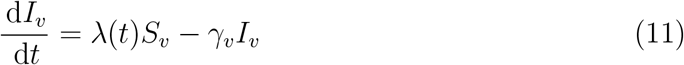

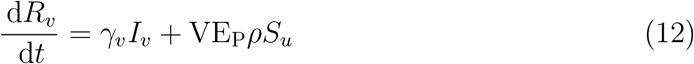

This is the approach used in “status-based” models of cross immunity—such models keep track of immune statuses of individuals, rather than their infection histories (Gog and Grenfell, 2002; Gog and Swinton, 2002; Kucharski et al., 2016). For this model, the parameter VE_P_ is the measure of vaccine efficacy.

These two widely used models have important dynamical differences. For a given set of shared parameters, and the same value of vaccine efficacy, initial dynamics will be the same, but the permanent protection of individuals in the polarized model will always result in a lower final outbreak size than the leaky vaccination model. When both VE and the initial value of *ℜ* are relatively high, this difference is large.

To better understand this gap, we consider an immune-boosting model. The leaky vaccination model assumes that vaccinated individuals are challenged with a lower force of infection (1− VE_L_)*λ*(*t*), but in general it is not realistic to assume that challenges would completely disappear only because of immune status. In a homogeneously mixing population, we expect both vaccinated and unvaccinated individuals to be challenged with identical forces of infection *λ*. Therefore, the leaky vaccination model implicitly assumes that vaccinated individuals have an *independent* probability (1 − VE_L_) of infection for every challenge. Instead, the immune-boosting model assumes that unsuccessful challenges elicit immune response, moving individuals from *S*_*v*_ to *R*_*v*_ compartment at rate VE_L_*λ*(*t*) and thereby breaking the independence assumption of the leaky vaccine model:

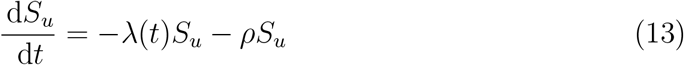

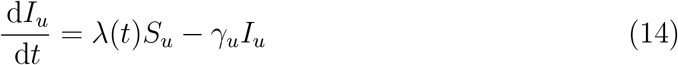

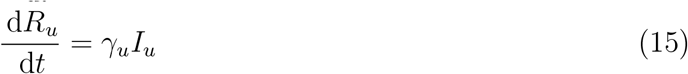

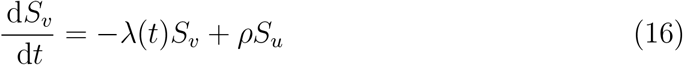

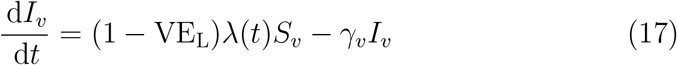

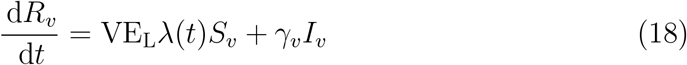

In this model, both unvaccinated and vaccinated individuals are subject to identical forces of infection, which represent the per capita rate of challenges, but the outcome of challenges differ.

The epidemiological dynamics (i.e., trajectories of *I*_*u*_ and *I*_*v*_) predicted by the immune-boosting model (based on leaky vaccination) and the polarized vaccination model are identical: both models assume that individuals become vaccinated at rate *ρ* and move out of the *S*_*v*_ compartment at rate *λ* and only differ in when individuals get sorted based on the result of their next challenge. This equivalence allows us to bridge the difference between the leaky and polarized vaccination models. The equivalence holds regardless of infection characteristics of vaccinated individuals (i.e., the duration of their infection and their transmissibility). In Supplementary Materials, we further show that epidemic dynamics are independent of the shape of the susceptibility distribution under immune boosting (and instead only depends on the mean susceptibility); under a leaky vaccination model, however, epidemic dynamics are sensitive to the susceptibility distribution (Gomes et al., 2014).

Finally, we consider a generalized model that encompasses all three mechanisms above (dichotomous vaccine responses, partial protection, and immune boosting):

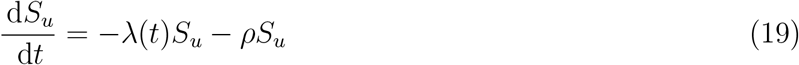

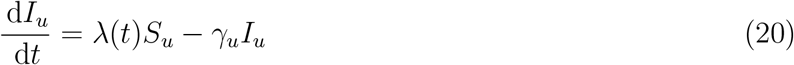

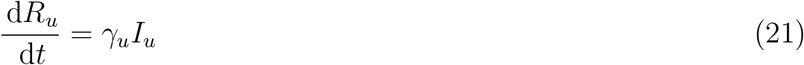

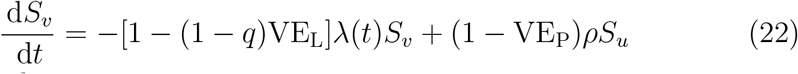

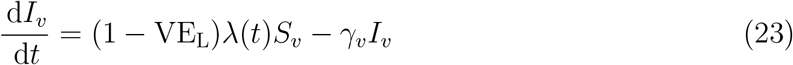

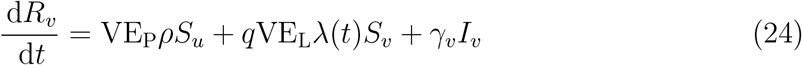

This model includes one new parameter, *q*, which represents the proportion of unsuccessful challenges that result in immune boosting. When *q* = 0 (i.e., in the absence of boosting), setting VE_P_ = 0 gives us the leaky vaccination model. When *q* = 1 (i.e., in the presence of full boosting), setting VE_P_ = 0 gives us the immune-boosting model, whereas setting VE_L_ = 0 gives us the polarized vaccination model. The relationship between these four models are summarized in Fig. 1. The generalized vaccination model has a combined vaccine efficacy of VE = 1 (1 VE_L_)(1 VE_P_). We later analyze the dynamics of the generalized vaccination model while keeping VE fixed.

**Figure 1.**
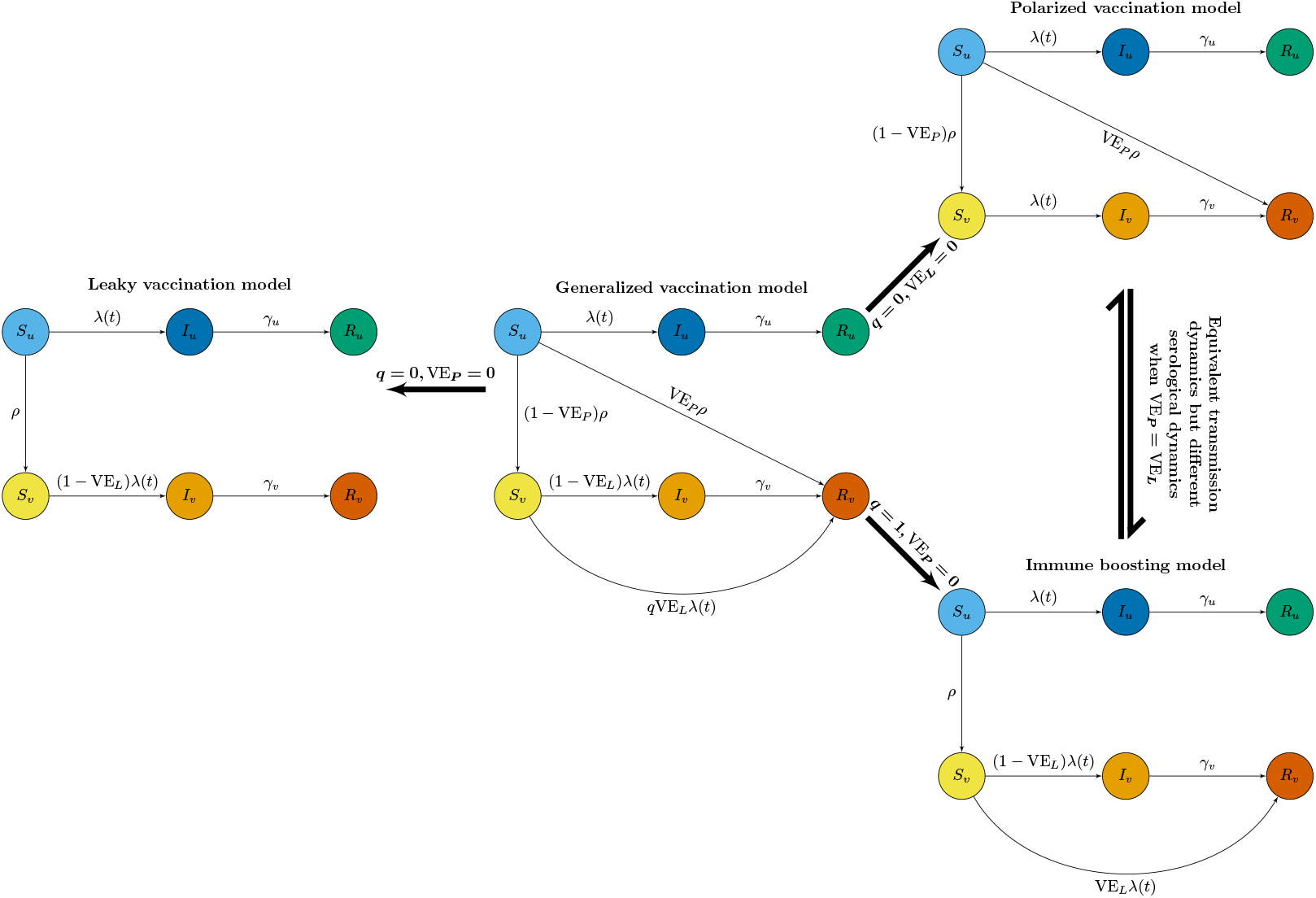
A schematic diagram of four different vaccination models. *S* represents susceptible individuals. *I* represents infected individuals. *R* represents recovered individuals. *λ* represents force of infection. *ρ* represents the rate of vaccination. *p* represents vaccine efficacy. *γ* represents recovery rate. *θ* represents the proportion of individuals that remain partially susceptible after vaccination. *q* represents the proportion of unsuccessful challenges that result in immune boosting. Subscripts *u* and *v* represents unvaccinated and vaccinated.

## Model simulations

We begin by comparing the dynamics of three individual models: leaky vaccination, polarized vaccination, and immune-boosting models. As an example, we consider a homogeneously mixing population. In this case, the force of infection is given by:

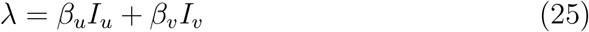

For simplicity, we assume that, once infected, both unvaccinated and vaccinated individuals transmit at the same rate *β*_*u*_ = *β*_*v*_ = 0.5*/*day for an average of 1*/γ* = 5 days. We also assume that *φ* = 0.5 proportion of individuals are vaccinated at the beginning of an epidemic with 60% efficacy (VE_P_ or VE_L_ = 0.6) and that vaccination does not continue during the out-break (*ρ* = 0). For the leaky vaccination model and the immune-boosting model, we set *S*_*v*_(0) = 1 − *φ* and *R*_*v*_(0) = *φ*. For consistentency, we then set *S*_*v*_(0) = *φ*(1 *−*VE_P_) and *R*_*v*_(0) = *φ*VE_P_ as our initial condition for the polarized vaccination model.

Fig. 2 compares epidemiological (A–C) and immune-status (D–F) trajectories predicted by the three models. As explained earlier, the leaky vaccination model predicts more infections among vaccinated individuals than the other two models, which predict identical incidence trajectories. The leaky vaccination model also predicts more among unvaccinated individuals because a larger outbreak among vaccinated individuals causes unvaccinated individuals to experience a greater forces of infection over time.

**Figure 2.**
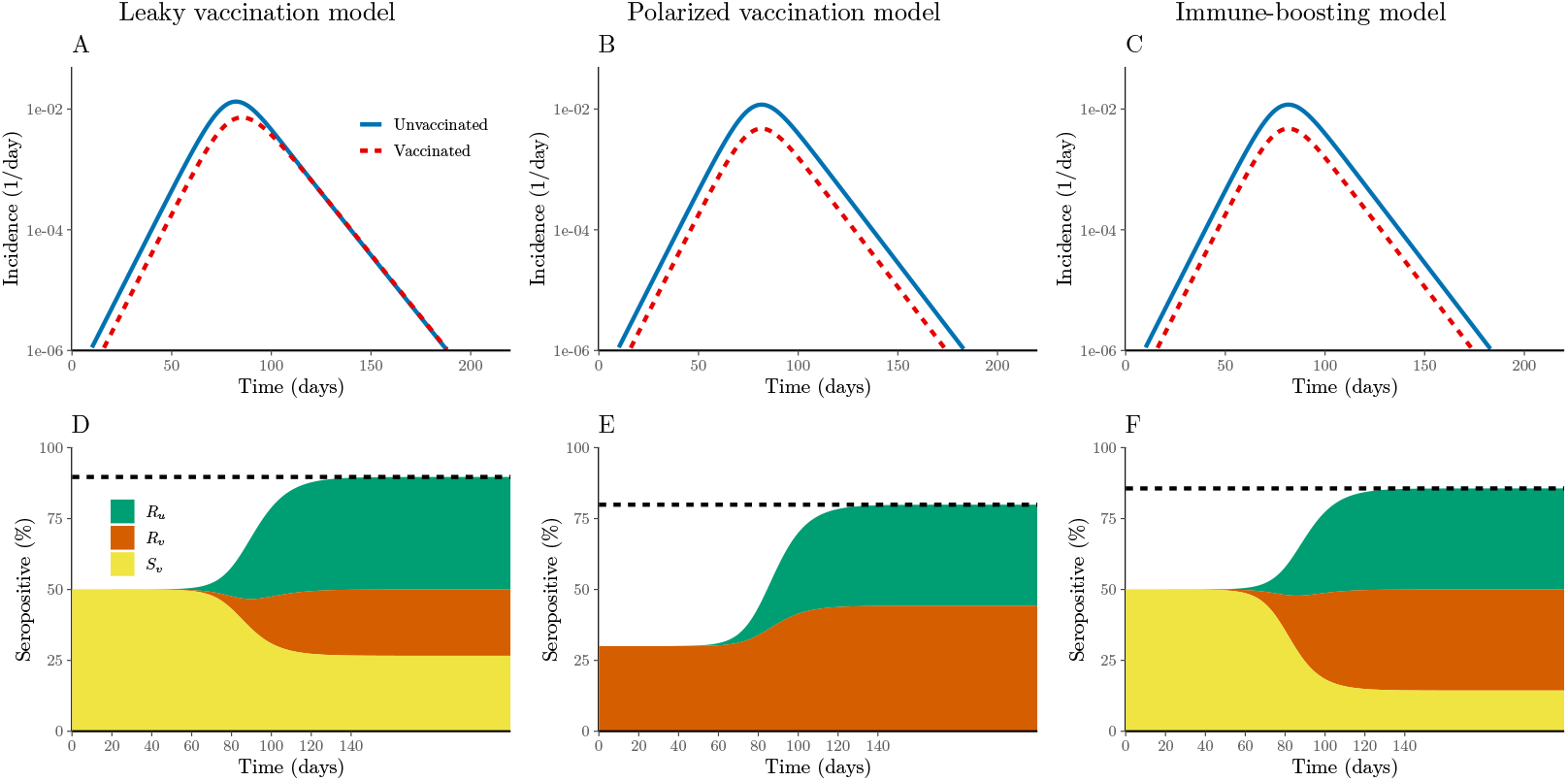
Simulations of three different vaccination models. (A–C) Incidence of infection among unvaccinated (blue solid) and vaccinated (red dashed) individuals. (D–F) Immune status over time (compartments *R*_*u*_, *S*_*v*_, and *R*_*v*_). The *S*_*v*_ compartment is not included in the polarized vaccination model because it represents a set of individuals who have not retained any immunity from vaccination. Simulations are performed assuming *β*_*u*_ = *β*_*v*_ = 0.5*/*day for an average infectious periods of 1*/γ* = 5 days. We also assume that *φ* = 0.5 proportion of individuals are vaccinated at the beginning of an epidemic with 60% efficacy (VE_P_ = VE_L_ = 0.6) and that vaccination does not continue during the outbreak (*ρ* = 0).

We further find that all three models predict different immune-status trajectories. (Fig. 2D–F). Here, we do not distinguish the sources of antibodies (whether derived from natural infections or vaccinations) and assume that individuals in *R*_*u*_, *S*_*v*_, and *R*_*v*_ compartments are seropositive, except in the case of polarized vaccination: in such case, we assume individuals in the *S*_*v*_ compartment are seronegative because they have not retained any immunity from the vaccination. The leaky vaccination model predicts the largest out-break and therefore the highest levels of seroprevalence (89.7% by the end of the simulation). The immune-boosting model predicts lower seroprevalence (85.6%), reflecting the lower final size, while the polarized vaccination model predicts a still lower seroprevalence (79.9%) because of our assumption that people not protected by polarized vaccination do not are not seropositive.

We next use the generalized vaccination model to further investigate how the final size of the an epidemic among vaccinated individuals depends on assumptions about vaccine-derived immunity across a wide range of assumptions about the basic reproduction number *ℜ*_0_ and vaccine efficacy VE (Fig. 3). In particular, we factor vaccine efficacy VE in terms of leaky vaccine efficacy VE_L_ and polarized vaccine efficac VE_P_, and consider an intermediate case, in which 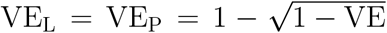, as well as the extreme cases, in which case VE_L_ = VE or VE_P_ = VE. First, when VE_L_ = VE, all vaccinated individuals have identical susceptibility; in this case, increasing the amount of boosting *q* reduces the final size as expected (see first column of Fig. 3). We observe biggest effects of boosting at intermediate vaccine efficacy, VE, and high basic reproduction number, *ℜ*_0_ (see bottom left panel of Fig. 3). When vaccine efficacy is too low (or too high), then boosting has negligible effects because virtually everyone (or virtually no one) gets infected. As we increase *ℜ*_0_, the leaky vaccination model predicts that all vaccinated individuals will eventually get infected. On the other hand, the final size predicted by the immune-boosting model cannot be greater than 1 − VE. As we increase VE_P_ (and decrease VE_L_ accordingly), the generalized vaccination model collapses to the polarized vaccination model, and the final size becomes insensitive to the boosting parameter *q*.

**Figure 3.**
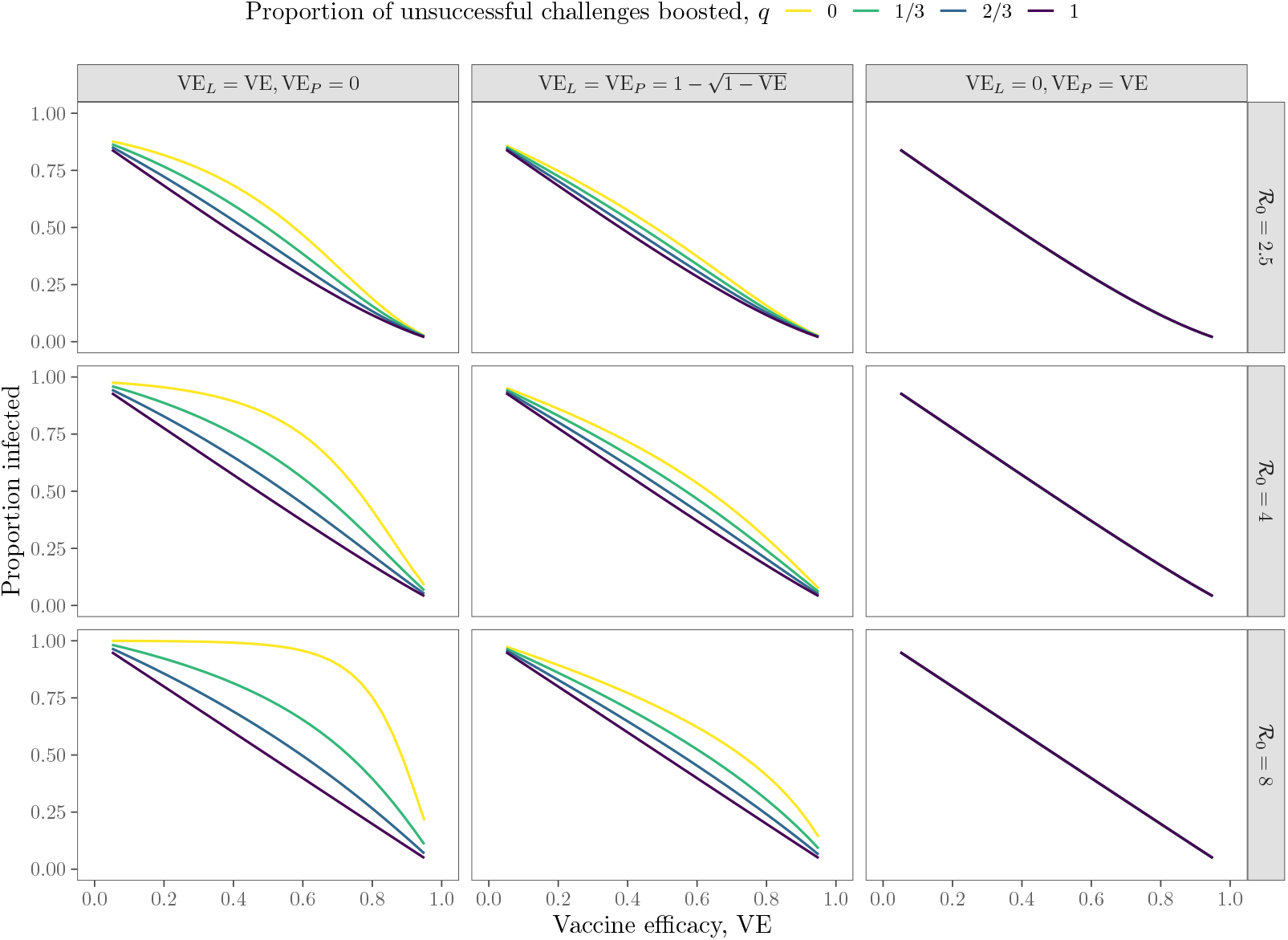
Sensitivity of the final size of an outbreak among vaccinated individuals to assumptions about vaccine-derived immunity. Final size of an outbreak was calculated by simulating the generalized vaccination model for 220 days. All other parameters are the same as in Fig. 2.

So far, we have limited our discussions to vaccine efficacy, which we defined as the proportion of people protected from their first challenge. We distinguish this from vaccine *effectiveness*, which is measured empirically (Halloran et al., 2009). Here, we compare two ways of estimating vaccine effectiveness: using cumulative incidence or instantaneous hazard. Several factors can cause vaccine effectiveness to systematically differ from vaccine efficacy—in our case, the main reason is the fact that some vaccinated individuals may be challenged multiple times.

Cumulative incidence refers to the cumulative proportion of infections among unvaccinated and vaccinated individuals; this is commonly used for measuring the vaccine effectiveness in real outbreaks (Farrington, 1993). Since we are modeling a single epidemic without a loss of immunity or multiple infections, we consider the reduction in cumulative incidence throughout the entire epidemic. To do so, we add two additional compartments, which keep track of cumulative incidence among unvaccinated *C*_*u*_ and vaccinated

*C*_*v*_ individuals:

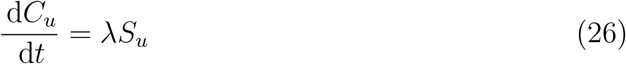

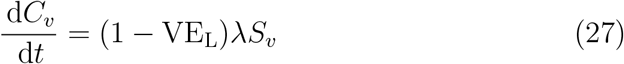

Since we are neglecting vaccinations that occur during the outbreak (*ρ* = 0, the cumulative proportion of infections among vaccinated *p*_*v*_(*t*) and unvaccinated *p*_*u*_(*t*) individuals can be expressed as:

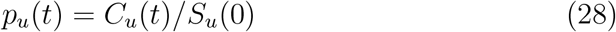

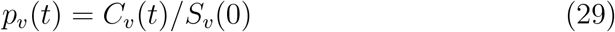

Then, the estimated vaccine effectiveness at time *t* is:

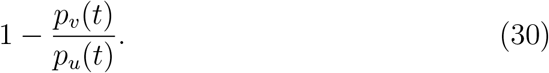

On the other hand, instantaneous hazard refers to the per-capita rate at which unvaccinated *h*_*u*_(*t*) and vaccinated *h*_*v*_(*t*) individuals get infected if they have not yet been infected yet. These quantities can be calculated by dividing the incidence of new infection by the number of uninfected individuals. The per-capita rate of infection *h*_*v*_(*t*) among vaccinated individuals in then given by:

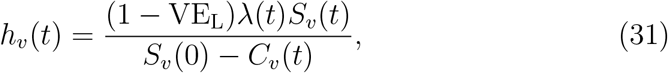

where *S*_*v*_(0) − *C*_*v*_(*t*) *≥S*_*v*_(*t*) because vaccinated individuals can leave the *S*_*v*_(*t*) compartment via boosting; in other words, we are assuming that boosting is not observed, and that boosted individuals are neither counted as infected, nor removed from the denominator. The per-capita rate of infection *h*_*u*_(*t*) among unvaccinated individuals is straightforward:

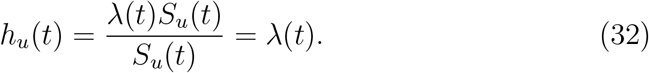

Then, the estimated reduction in hazard at time *t* is:

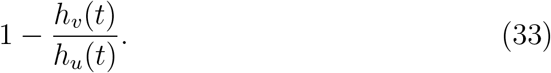

We compare two estimates of vaccine effectiveness across a wide range of assumptions about vaccine-derived immunity in Fig. 4. We assume 60% efficacy throughout (therefore VE = 0.6). Under polarized vaccination (VE_P_ = VE, VE_L_ = 0), the cumulative-incidence reduction always gives correct answers throughout the epidemic—since the susceptible pool among unvaccinated and vaccinated individuals is depleted at the same rate *λ*, the ratios of their proportions of cumulative infections remain constant. Likewise, the cumulative-incidence reduction also gives correct answers under immune boosting (*q* = 1).

**Figure 4.**
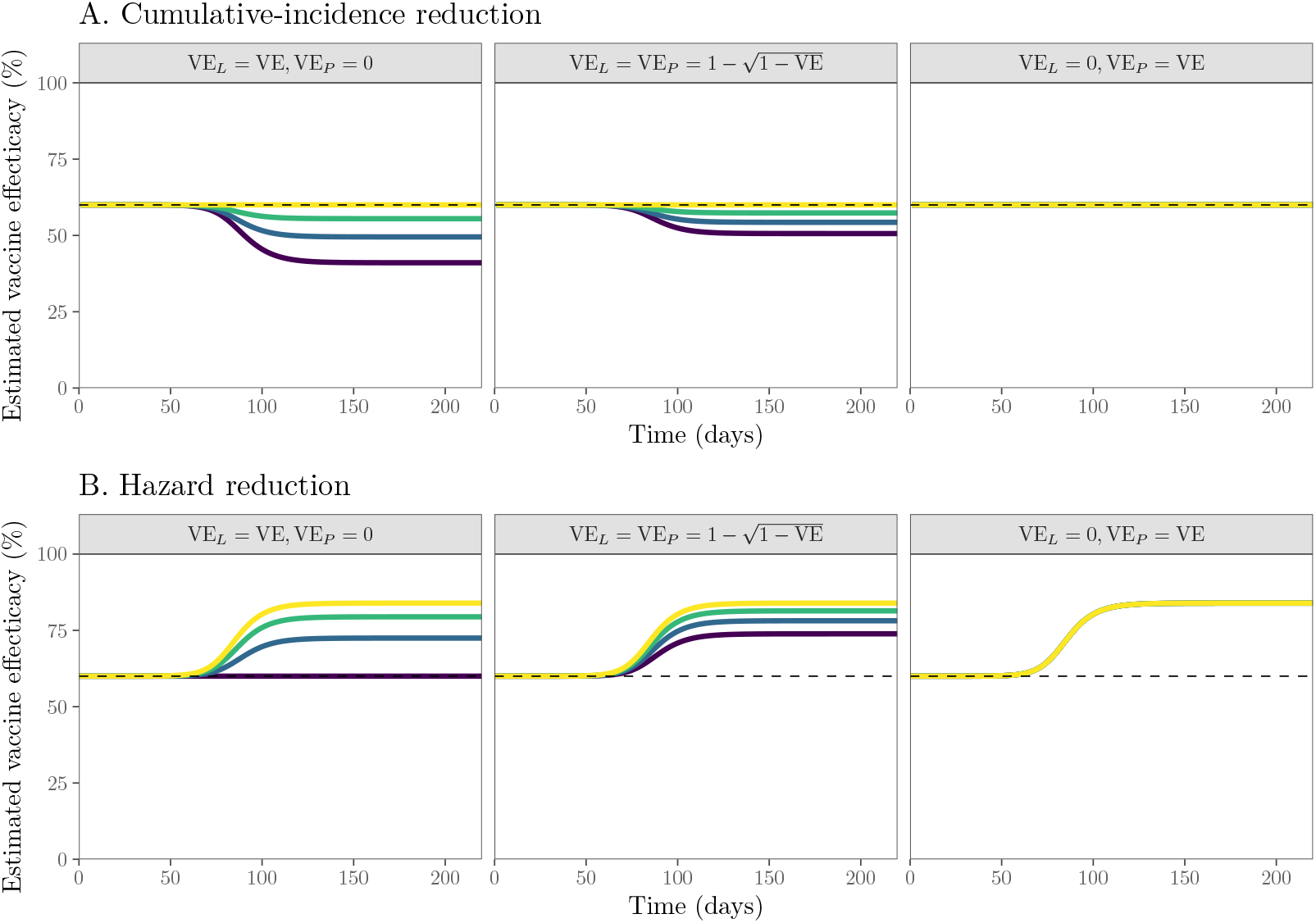
Estimates of vaccine effectiveness using reduction in cumulative incidence (A) and hazard (B) over time. Vaccine effectiveness was calculated by simulating the generalized vaccination model for 220 days. Colored lines represent the estimated vaccine effectiveness. Dashed lines represent the assumed vaccine efficacy. We assume *ℜ*_0_ = 2.5 and a combined efficacy of VE = 0.6 throughout. All other parameters are the same as in Fig. 2.

Likewise, the cumulative-incidence reduction also give correct answers for the polarized vaccination model (VE_L_ = 0, VE_P_ = VE). However, when some challenges are not boosted (*q <* 1), using cumulative incidence underestimates the vaccine efficacy beyond the exponential growth phase. This is because vaccinated individuals who have been exposed but are not boosted or infected still remain susceptible to future infections; larger final epidemic sizes predicted by these models (Fig. 3) then translate to a seemingly lower vaccine efficacy.

The hazard reduction gives correct answers for the leaky vaccine model (when *q* = 0, VE_L_ = VE, and VE_P_ = 0) because the ratios of force of infection that unvaccinated and vaccinated individuals experience are always constant. However, the hazard reduction overestimates vaccine efficacy in the presence of immune boosting: since boosted individuals have not yet been infected, the susceptible pool in the vaccinated group appears to be bigger than it really is, causing the per-capita rate of infection to seem smaller. Vaccine efficacy is also overestimated for polarized vaccination for similar reasons.

We note that both estimates give correct answers during the exponential growth phase, regardless of underlying assumptions about immunity. More generally, we expect both estimates to give unbiased estimates as long as the depletion of susceptible pool is negligible among both vaccinated and unvaccinated individuals; in trial settings, where incidence is relatively low, this assumption may hold. But estimating vaccine effectiveness from real outbreaks is expected to be more difficult.

## Discussion

Understanding the degree to which vaccination provides protection against infections is critical to predicting epidemic dynamics. The polarized model has been largely neglected in epidemiological modeling, in part due to its apparently extreme assumption that a fraction of vaccinated individuals do not receive any protection. But the leaky vaccination model also makes an unrealistic assumption: that vaccinated individuals who are exposed to infections can still remain susceptible, independent of previous exposures. This assumption causes the leaky vaccination model to always predict a larger epidemic final size. This difference can be bridged with immune boosting. With boosting, vaccinated individuals can attain protection without developing a transmissible infection. In particular, the leaky model with perfect immune-boosting model predicts identical epidemic dynamics to the polarized vaccination model because individuals in both cases are completely immune after surviving a single challenge.

Even though immune boosting and polarized vaccination models predict the same epidemic dynamics, they may have different immune-status dynamics. We investigate both aspects using a generalized vaccination model, which encompasses the mechanisms of all three models. The generalized vaccination model includes one additional parameter, which determines the amount of immune boosting. We use this model to show that the epidemic dynamics are most sensitive to the assumptions about vaccine-derived immunity at an intermediate vaccine efficacy.

Finally, assumptions about vaccine-derived immunity also have important implications for estimating vaccine effectiveness. Vaccine effectiveness can be estimated based either on cumulative incidence or on hazard rates. Cumulative-incidence-based effectiveness estimates will reflect initial efficacy for polarized vaccination and immune-boosting models, whereas hazard-based estimates reflect efficacy for the leaky vaccination model. Neither method reflects efficacy for intermediate cases. These differences are driven by different assumptions about what happens when individuals are challenged more than once; thus both methods reflect efficacy when the cumulative hazard of infection is low. Conversely, interpretation of effectiveness estimates when a large fraction of unvaccinated individuals have been infected depends on (usually unknown) details of immune dynamics.

We rely above on a simplifying assumption that natural infections (as well as polarized vaccination and immune boosting) provide permanent protection against future infections. In practice, both infection- and vaccine-derived immunity wane over time for many pathogens (Heffernan and Keeling, 2009; Lewnard and Grad, 2018; Pérez-Alós et al., 2022). When immunity wanes, polarized vaccination and immune-boosting models may not necessarily predict identical dynamics. In particular, individuals who gain complete protection through polarized immunity may immediately enter the *R*_*v*_ compartment upon vaccination, whereas those who gain complete protection through immune boosting take longer to enter the *R*_*v*_ compartment because they need to be exposed to infections. These differences can translate to shorter delays between reinfection events for the polarized immunity model, which in turn can lead to dynamical differences at the population level.

There are also other complexities that need to be considered. For example, individuals who are boosted after vaccination can have different immunity profiles compared to those who attained strong protection from vaccination alone. These individuals also likely have different immunity profiles from those who have been infected but never been vaccinated. These differences can also cause polarized vaccination and immune-boosting models to behave differently. Despite these limitations, immune boosting, which is often neglected in epidemic models of vaccination, is still expected to be an important mechanism for understanding dynamics of many pathogens.

We have provided a unifying framework for understanding the impact of vaccination on the spread of infectious disease. The specifics of how vaccination translates into immunization defines the population burden of infection via its effect on the epidemic final size. Yet discussion of how the two extreme models commonly used (leaky and polarized) are related has been lacking. By making this link, we both illustrate the spectrum of trajectories expected for a range of configurations, and illuminate the effects of these assumptions on medium-term vaccine effectiveness.

## Data Availability

All data and code are stored in a publicly available GitHub repository (https://github.com/parksw3/immune-boosting).

## Supplementary Text

Here, we show that, in the presence of immune boosting, epidemic dynamics are independent of the shape of the susceptibility distribution (depending only on mean susceptibility). To do so, consider an immune-boosting model that allows for heterogeneity in vaccine-derived immunity. We assume that a vaccinated individual’s susceptibility 0 *≤p≤* 1 follows some distribution *f* (*p*):

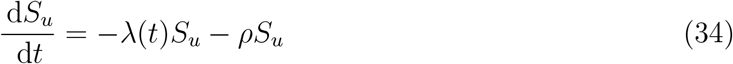

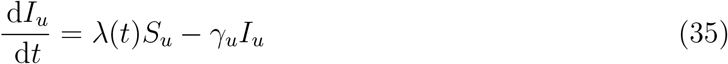

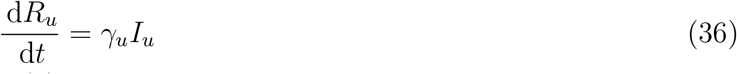

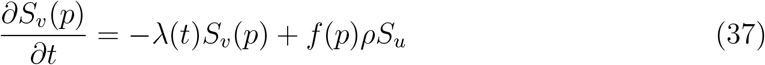

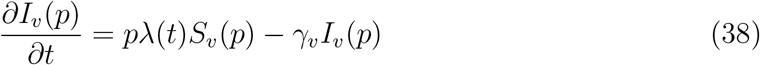

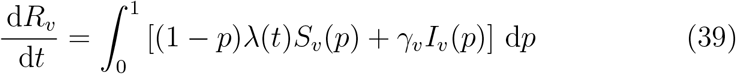

Due to immune boosting, *S*_*v*_(*p*) is always depleted at a per-capita rate of *λ*(*t*) regardless of the values of *p*, meaning that the (normalized) distribution of *S*_*v*_(*p*) will always follow *f* (*p*). To obtain the dynamics of total prevalence *I*_*v*_ = *I*_*v*_(*p*) d*p*, we can integrate *∂I*_*v*_(*p*)*/∂t* across *p*:

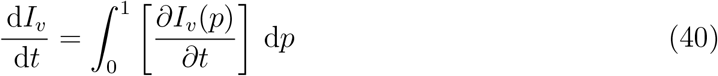

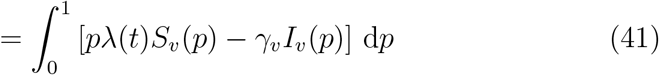

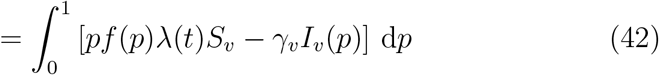

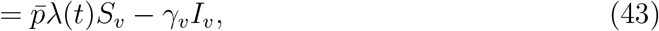

where 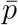 represents the mean of the distribution *f* (*p*), and *S*_*v*_ = *S*_*v*_(*p*) d*p* represents the proportion of total susceptible, vaccinated individuals. Therefore, the dynamics of total prevalence *I*_*v*_ depends only on the mean sus-ceptibility boosting. 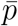 and not on the shape of the distribution *f* (*p*) under immune

